# Benefits of integrated screening and vaccination for infection control

**DOI:** 10.1101/2021.12.18.21268047

**Authors:** Marie Jeanne Rabil, Sait Tunc, Douglas R. Bish, Ebru K. Bish

## Abstract

**Importance:** Screening and vaccination are essential in the fight against infectious diseases, but need to be integrated and customized based on community and disease characteristics.

**Objective:** To develop effective screening and vaccination strategies, customized for a college campus, to reduce COVID-19 infections, hospitalizations, deaths, and peak hospitalizations.

**Design, Setting, and Participants:** We construct a compartmental model of disease spread for vaccination and routine screening, and study the efficacy of four mitigation strategies (routine screening only, vaccination only, vaccination with partial routine screening, vaccination with full routine screening), and a no-intervention strategy. The study setting is a hypothetical college campus of 5,000 students and 455 faculty members, with 11 undetected, asymptotic SARS-CoV-2 infections at the start of an 80-day semester. For sensitivity analysis, we vary the screening frequency, daily vaccination rate, initial vaccination coverage, and screening and vaccination compliance; and consider three scenarios that represent low/medium/high transmission rates and test efficacy. Model parameters come from publicly available or published sources.

**Results:** With low initial vaccination coverage, even aggressive vaccination and screening result in a high number of infections: 1,024/2,040 (1,532/1,773) with routine daily (every other day) screening of the unvaccinated; 275/895 with daily screening extended to the newly vaccinated in base- and worst-case scenarios, with reproduction numbers 4.75 and 6.75, respectively, representative of COVID-19 Delta variant. With the emergence of the Omicron variant, the reproduction number may increase and/or effective vaccine coverage may decrease if a booster shot is needed to maximize vaccine efficacy.

**Conclusion:** Integrated vaccination and routine screening can allow for a safe opening of a college when initial vaccination coverage is sufficiently high. The interventions need to be customized considering the initial vaccination coverage, estimated compliance, screening and vaccination capacity, disease transmission and adverse outcome rates, and the number of infections/peak hospitalizations the college is willing to tolerate.

## Introduction

The current COVID-19 pandemic has illustrated how unprepared many countries were to combat such a highly contagious disease with potentially severe health outcomes. On the science side, screening kits, then vaccines, were developed at record speed, and their availability increased over time. On the implementation side, however, screening and vaccination efforts were initially administered on a mostly *ad hoc* basis, and more importantly, these efforts were initially uncoordinated.

First came the screening kits (around January 2020 [10]), making COVID-19 screening possible. Early on, the number of screening kits was limited [37, 50], and screening focused mainly on symptomatic individuals (with isolation of those detected with the infection). As availability increased, more routine screening, especially in residential facilities where people lived in close proximity to each other (e.g., college campuses, nursing homes, prisons, military bases), was deployed. Starting in December 2020 [17], vaccines started receiving emergency approval by the Food and Drug Administration (FDA) in the United States (U.S.), and by respective regulatory agencies in other parts of the world. As vaccine supplies increased, vaccines were progressively made available to the public based on pre-established priority groups. Initially these policies lacked coordination among screening and vaccination efforts, necessitating research to develop decision-making tools as well insights on effective, coordinated interventions. By developing such a tool and insights on effective vaccination and routine screening practices for college campuses, this paper contributes to this stream of research.

The literature on screening and vaccination for infectious diseases is vast, with many studies on the benefits of, and effective strategies for, lab-based screening, e.g. [1, 14, 34, 43, 56], and vaccination, e.g. [3, 22, 31, 35, 40]; and we refer the interested reader to the many references in these works. The study by [19] pairs antibody testing with vaccination, but does not consider routine screening. The studies by [29, 33] discuss routine screening on a college campus, but without on-campus vaccination efforts, that is, only considering that some portion of campus residents are already vaccinated prior to the start of the semester (“initial vaccine coverage”); in this setting, these studies investigate the impact of different levels of vaccine efficacy and initial vaccine coverage on the number of infections. The study by [61] discusses the combination of initial vaccine coverage, on-campus screening, and on-campus population size (e.g., 100% on-campus population size implies that all students take classes in person) to control outbreaks on a college campus.

We contribute to this stream of literature by building a detailed compartmental model to construct integrated on-campus vaccination and routine screening strategies for a college campus, and discuss how the effectiveness of the different strategies depend on community and disease characteristics, including initial vaccine coverage, estimated compliance for on-campus screening and vaccination, available screening and vaccination supplies/capacity, disease transmission and adverse outcome rates, as well as the number of infections and peak hospitalizations the college is willing to tolerate. In particular, our model is more detailed than those in the literature, and is built specifically to model strategies that not only include initial vaccine coverage, but also on-campus vaccination, as well as different variants of routine on-campus screening, to allow for the targeted screening population to include only the unvaccinated, or the unvaccinated and the newly vaccinated. We perform a detailed comparative study of the different interventions, characterized by the level of on-campus vaccination and screening, for various community and disease characteristics and disease-related outcome measures.

Various protective and/or preventative interventions, such as screening, vaccination, and isolation of those infected, can be employed to combat a pandemic. An optimal strategy may involve a combination of the different interventions, implemented with different frequencies, and needs to be revised over time and customized for each region, as the available interventions (e.g., screening kits, vaccine) and their supplies, and the local situation of the pandemic change. In this paper, we focus on integrated strategies of routine screening and vaccination, which we always pair with the isolation of infected individuals detected through screening. We show, through a case study of a college campus, that vaccination and screening work synergistically, and lower the total number of infections and hospitalizations, and the number of infections and hospitalizations at the peak day, thus flattening the infection curve, over what either intervention can achieve alone given an initial vaccine coverage; however, the effectiveness of such on-campus interventions also depend heavily on the initial vaccine coverage. In addition to the timely detection and quarantine of infected individuals, routine screening further allows for an efficient allocation of the limited vaccine supplies: It does so by removing those individuals who are infected and asymptomatic, or those with a prior infection, from the vaccination pool. This redirects on-campus vaccination efforts from those who would not receive much benefit, to those who would. On the other hand, vaccination shrinks the susceptible population. We also provide insight on strategies that are effective in various phases of a pandemic response, characterized by different availability of screening and vaccination supplies, current vaccine coverage, and the severity of infection spread (transmission rate) in the community, as well as adverse outcome rates of the infection.

## Methods

### Study Design and Parameters

We develop an extended compartmental epidemic model to study the spread of an infectious disease in a population composed of two groups (students and faculty/staff, hereafter students and faculty) under protective and/or preventative interventions, including screening, isolation, and vaccination; and utilize our model to study the spread of COVID-19 on a college campus. Our model provides an extension to an existing compartmental framework [34], to model integrated screening and vaccination strategies under various compliance levels, while capturing different disease and transmission dynamics of the faculty (indexed by “f”) and students (indexed by “s”). The details of the new compartmental model can be found in the S1 Appendix.

#### Strategy Description

We consider routine screening that excludes symptomatic individuals who go into isolation immediately upon the start of their symptoms (following a separate testing process, as needed). Regarding routine screening, the subjects to be screened each day are selected according to the given screening strategy (i.e., screening population and frequency). *Partial screening* applies to those subjects that are either susceptible, or “thought” to be susceptible; while *full screening* extends routine screening to “vaccinated-but-not-yet-immune” subjects. All screening uses the polymerase chain reaction (PCR) test, which can detect, with high accuracy, the presence of the virus. [26, 52] It takes 8 hours to receive the test result, and a positive test result leads to the immediate isolation of the affected subject. For vaccination, we consider that some proportion of each group starts the semester as fully vaccinated (i.e., with initial immunity). Then, subjects to be vaccinated each day on campus are randomly selected from those subjects that are either susceptible, or “thought” to be susceptible (thus excluding those initially immune), and is restricted by the given vaccination rate per day. We assume that the vaccines administered on campus correspond to either the two-dose Pfizer vaccine (60%) or the two-dose Moderna vaccine (40%) [46]. We do not consider one-dose vaccines, because they represent less than 4% of the total number of vaccines administered in the U.S. [46]; and we do not consider waning immunity for those vaccinated (either at the outset or while on campus) due to the relatively short, 80-day period of the semester. Overall, we consider four strategies that represent various combinations of vaccination and screening efforts, and the no-intervention strategy, as summarized in Table 1. We assume perfect compliance for all isolation orders, and model imperfect compliance for screening and vaccination.

**Table 1.** Strategy description (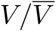 denotes vaccination/no vaccination; 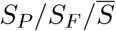 denotes partial/full/no screening)

| Strategy | Definition |
| --- | --- |
| Strategy $\bar{V} - \bar{S}$<br>(no intervention) | No vaccination<br>and no screening |
| Strategy $\bar{V} - S_F$ | No vaccination<br>and full screening |
| Strategy $V - \bar{S}$ | Vaccination<br>with no screening |
| Strategy $V - S_P$ | Vaccination<br>with partial screening |
| Strategy $V - S_F$ | Vaccination<br>with full screening |
| Sensitivity Analysis |  |
| Daily vaccination rate | 30/day, 60/day, 120/day |
| Screening frequency | 1 day, 2 days, 3 days, 7 days, 14 days |
| Initial vaccine coverage for students ($L_s$) | 30%, 60%, 90% |
| Initial vaccine coverage for faculty ($L_f$) | 30%, 60%, 90% |
| Vaccination compliance rate ($\alpha$) | 50%, 75%, 95% |
| Screening compliance rate ($\eta$) | 50%, 75%, 95% |
| Severity scenarios | worst-case, base-case, best-case |

#### Sensitivity Analysis

We conduct various sensitivity analyses through three scenarios, which represent different levels of pandemic severity and test efficacy, and through varying the values for the daily vaccination rate, screening frequency, compliance (for each of screening and vaccination), and initial vaccine coverage (for each of faculty and students). Each scenario is characterized by the level of infection spread (modeled by group transmission rates, *β*(*f*) and *β*(*s*), reported in terms of an overall reproduction number, *R*_*T*_, computed as a weighted average of each group’s reproduction number, see S1 Appendix); exogenous conditions (*X* weekly imported infections, i.e., *X* new infections introduced to campus residents each week from external contacts, such as family or community members); and test efficacy, in terms of test specificity (*spec*) and sensitivity (*sens*).

We label these scenarios as the *worst-case* (high infection spread, low test sensitivity and specificity, high imported infections), *best-case* (low infection spread, high test specificity, medium test sensitivity, low imported infections), and *base-case* (medium infection spread, low test specificity, medium test sensitivity, medium imported infections), Table 2. For each scenario, we vary the screening frequency (every 1,2,3,7, and 14 days) for all strategies that involve screening; and the daily vaccination rate (30/day, 60/day, 120/day) for all strategies that involve vaccination, Table 1. These strategies represent different situations where screening frequency and daily vaccination rate may be restricted due to limited testing capacity, testing kits, or vaccine supply; the proportion of students who are exempt from a vaccination mandate or are unwilling to be vaccinated; or other logistical issues involved with test or vaccine administration. Moreover, we consider different compliance rates for screening (*η*) and vaccination (*α*), Table D in S1 Appendix, and different levels of initial vaccine coverage for students (*L*_*s*_), Table B in S1 Appendix and faculty (*L*_*f*_), Table C in S1 Appendix, for the different strategies.

**Table 2.** Summary of key parameters

| Model Parameter | Value(s) | Comments/References |
| --- | --- | --- |
| **Disease dynamics** |  |  |
| Mean incubation time | 3 days | Paltiel et al. [34], 2020 |
| Time to recovery | 14 days | CDC [8], 2021 |
| Probability of symptoms if infected | 30% | Assumption (similar to Paltiel et al. [34], 2020), also based on Poletti et al. [36], 2021 |
| Fatality rate (students) | 0.05% | See S1 Appendix for calculations |
| Fatality rate (faculty) | 2% | Statista [45, 47], 2021. See S1 Appendix for calculations |
| Hospitalization rate (students) | 1.4% | CDC [6], 2021 and COVID-Net [13], 2021. See S1 Appendix for calculations |
| Hospitalization rate (faculty) | 8.4% | CDC [6], 2021 and COVID-Net [13], 2021. See S1 Appendix for calculations |
| Population reproduction number ($R_T$) | 4.75, 5.75, 6.75 | See S1 Appendix for calculations |
| Exogenous infections ($X$) | 5, 10, 25 per week | Assumption (similar to Paltiel et al. [34], 2020) |
| **Screening test characteristics** |  |  |
| Test sensitivity (*sens*) | 70%<br>80% | Based on Kortela et al. [23], 2021<br>Based on Stites & Wilen [48], 2020 and Woloshin et al. [58], 2020 and Yohe [59], 2020 |
| Test specificity (*spec*) | 98% , 99.7% | Yohe [59], 2020, Stites & Wilen [48], 2020 |
| Time to false-positive return | 1 day | Assumption (similar to Paltiel et al. [34], 2020) |
| **Vaccine characteristics** |  |  |
| Vaccine efficacy ($\epsilon$) | 94.64% | See S1 Appendix for calculations |
| Time to reach full immunity | 5.4 weeks | See S1 Appendix for calculations |
| **Severity scenarios** |  |  |
| Scenario | Parameter Values | Comments/References |
| Base-case scenario | $R_T = 5.75$, $X = 10/\text{week}$,<br>$sens = 80\%$, $spec = 98\%$ | See comments/references above |
| Best-case scenario | $R_T = 4.75$, $X = 5/\text{week}$,<br>$sens = 80\%$, $spec = 99.7\%$ | See comments/references above |
| Worst-case scenario | $R_T = 6.75$, $X = 25/\text{week}$,<br>$sens = 70\%$, $spec = 98\%$ | See comments/references above |

#### Setting and Parameters

We consider a hypothetical college campus of 5,000 students and 455 faculty members, with 10 students and one faculty member having undetected, asymptotic SARS-CoV-2 infections at the outset (representing 0.2% of each group), and with some individuals arriving to campus as fully vaccinated (i.e., with initial immunity), over an 80-day academic semester. We consider the COVID-19 PCR test, which is the primary test used to detect SARS-CoV-2 [11], with sensitivity 70% to 80%, and specificity 98% to 99.7%, based on published works [23, 48, 58, 59]. Vaccination rates of 60/day or higher lead to the ability to vaccinate all the eligible subjects within the 80-day period (i.e., by the end of the semester) under all scenarios, assuming initial vaccine coverage of at least 30% for both faculty and students. Table 2 reports all key parameters, along with references, see S1 Appendix for details.

#### Outcome Measures of Interest

Total number of infections, hospitalizations, and deaths over the 80-day semester for each of the student and faculty groups; the peak number and peak day of new infections, the peak number of hospitalizations.

### Compartmental Model

We have expanded the SEIR-based (Susceptible, Exposed, Infectious, Removed) compartmental model of a published study [34], to study the integrated screening and vaccination strategies in Table 1 under various compliance levels, for a campus population consisting of two groups (faculty and students), with group-specific disease and transmission dynamics and initial vaccination coverage. In particular, our model contains additional compartments to capture the details of screening and vaccination, when applied, as well as the characteristics of the faculty and student groups, and has the following distinct features:

- We model that some individuals start the semester as fully vaccinated (i.e., with initial immunity). For the given daily vaccination rate, the subjects to be vaccinated are randomly selected from the unvaccinated and susceptible population (i.e., the “uninfected,” “exposed,” “asymptomatic,” and “recovered & unknown” compartments), and excludes those unvaccinated individuals whose infection is detected through routine screening (i.e., the “knowingly immune” compartment, as explained below) for all strategies that involve routine screening. However, due to imperfect compliance, not all available vaccines will be used on a given day, leading to waste.
- We consider a two-dose vaccine with 94.64% efficacy upon the full dose. The time to reach full immunity is stochastic with a mean of 5.4 weeks from the day of the first dose (see S1 Appendix for calculations), before which the subject has no immunity. Hence, vaccinated subjects can be infected before reaching full immunity. This modeling necessitates additional compartments for the vaccinated-but-not-yet-immune subjects, namely the “vaccinated-uninfected,” “vaccinated-exposed,” and “vaccinated-asymptomatic” compartments.
- Each day, routine screening is offered to everyone eligible for screening on that day according to the given screening frequency and the screening strategy utilized (i.e., partial or full screening). Due to imperfect compliance, however, not all selected subjects will undergo screening. The test has imperfect sensitivity and specificity (Table 2); and our model contains additional compartments to account for false-positives.
- A subject with a positive test result during routine screening can be either a “false-positive,” or “asymptomatic & infected.” It takes 8 hours to receive the test result, and any subject with a positive test result moves to isolation immediately upon receiving the test result, where no transmission can occur (similar to a published study [34]). All false-positives are corrected after being in isolation for one day (thus leave isolation), while both the “asymptomatic & infected” and “symptomatic & infected” subjects have an isolation time with a mean of 14 days, after which they move to the “knowingly immune” compartment, unless the symptomatic subjects are hospitalized before. We assume sufficient isolation capacity and perfect compliance for isolation.
- We differentiate between subjects that are knowingly vs unknowingly immune, via two new compartments: The “knowingly immune” compartment is for those subjects who gained immunity through either vaccination (at the outset or while on campus), a prior asymptomatic infection that was verified by a positive routine test result (leading to isolation), or a prior symptomatic infection (leading to isolation); and the “recovered & unknown” compartment is for those subjects who gained immunity through a prior asymptomatic infection that was not tested (hence not detected), i.e., these individuals became immune without their knowledge. As a result, the subjects in the “recovered & unknown” compartment cannot be differentiated from “uninfected” individuals, and hence they might get vaccinated or tested.

### Statistical Analysis

We implement the compartmental model in C++. The results are extracted to Microsoft Excel, which is used to plot different curves for the purpose of visualizing the results. No statistical tests are run, hence we do not report any statistical significance levels.

## Results

### Integrating Screening with Vaccination

In our hypothetical college campus of 5,000 students and 455 faculty members, 10 students and one faculty member have undetected, asymptotic SARS-CoV-2 infections (0.2% of each group) at the start of the semester.

We first discuss our results for the base-case scenario, considering 60% initial vaccination coverage for both students and faculty, and 75% compliance for both on-campus vaccination and screening. In this setting, vaccination alone (strategy 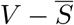), at a rate of 60/day, results in 1, 200 infections (1,185 students, 15 faculty) and a daily peak of 5 hospitalizations, while full routine screening alone (strategy 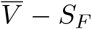), with an 80% sensitive and 98% specific test, results in 1, 408 infections (1,392 students, 16 faculty) if conducted weekly, and 294 infections (292 students, 2 faculty) if conducted daily, compared to the no-intervention strategy (strategy 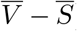) that results in 1, 834 infections (1,802 students, 32 faculty) and a daily peak of 8 hospitalizations over the 80-day semester in the base-case scenario. Augmenting the 60/day vaccination with routine screening of the unvaccinated population (strategy *V − S*_*P*_) every 1, 2, 3, 7, and 14 days results in 337, 513, 633, 880, and 1,024 infections, respectively, and 2 to 4 peak hospitalizations (Table 3). Including the vaccinated-but-not-yet-immune population in routine screening (strategy *V − S*_*F*_) further diminishes the number of infections (e.g., from 880 to 711 for weekly screening). When the vaccination rate increases (reduces) to 120/day (30/day), vaccination alone results in 448 (1,561) infections, and a peak of 1 (7) hospitalization(s) in the base-case. Augmenting the 30/day vaccination with routine screening of the unvaccinated population (strategy *V − S*_*P*_) every 1, 2, 3, 7, and 14 days results in 484 to 1,399 infections, and 3 to 6 peak hospitalizations (Table 3). All strategies with 120/day vaccination, and most strategies with 60/day vaccination lead to zero fatality, but strategies with 30/day vaccination or no vaccination lead to 1-2 deaths (per our campus of 5,455), highlighting the importance of vaccination in preventing deaths.

**Table 3.** Peak number, peak day, and number (students, faculty, overall) of infections, number of deaths, and peak and total number of hospitalizations (students, faculty, overall) over the 80-day semester for all strategies, implemented at various screening frequencies and vaccination rates, under the base-case scenario, considering 75% compliance (*η* = *α* = 75%) and 60% initial campus-wide immunity (*L*_*s*_ = *L*_*f*_ = 60%).

|  |  | Infections |  |  |  |  | Deaths | Hospitalizations |  |  |  |
| --- | --- | --- | --- | --- | --- | --- | --- | --- | --- | --- | --- |
| Strategy | Test Frequency | Peak number | Peak day | Student number | Faculty number | Total number | Total number | Peak number | Student number | Faculty number | Total number |
| **120 vaccines/day** |  |  |  |  |  |  |  |  |  |  |  |
| $V - S_F$ | every 14d | 9 | 25 | 344 | 3 | 347 | 0 | 1 | 1 | 1 | 2 |
|  | every 7d | 7 | 25 | 278 | 3 | 281 | 0 | 1 | 1 | 1 | 2 |
|  | every 3 d | 4 | 23 | 169 | 2 | 171 | 0 | 1 | 0 | 1 | 1 |
|  | every 2d | 3 | 16 | 124 | 1 | 125 | 0 | 1 | 0 | 1 | 1 |
|  | every 1d | 2 | 16 | 65 | 0 | 65 | 0 | 1 | 0 | 1 | 1 |
| $V - S_P$ | every 14d | 11 | 25 | 377 | 4 | 381 | 0 | 1 | 1 | 1 | 2 |
|  | every 7d | 10 | 25 | 333 | 3 | 336 | 0 | 1 | 1 | 1 | 2 |
|  | every 3d | 8 | 25 | 247 | 3 | 250 | 0 | 1 | 1 | 1 | 2 |
|  | every 2d | 7 | 25 | 205 | 2 | 207 | 0 | 1 | 0 | 1 | 1 |
|  | every 1d | 5 | 25 | 142 | 1 | 143 | 0 | 1 | 0 | 1 | 1 |
| $V - \bar{S}$ | N/A | 12 | 25 | 444 | 4 | 448 | 0 | 1 | 1 | 1 | 2 |
| **60 vaccines/day** |  |  |  |  |  |  |  |  |  |  |  |
| $V - S_F$ | every 14d | 11 | 30 | 914 | 10 | 924 | 1 | 4 | 5 | 3 | 8 |
|  | every 7d | 8 | 24 | 704 | 7 | 711 | 0 | 3 | 4 | 3 | 7 |
|  | every 3 d | 5 | 24 | 388 | 4 | 392 | 0 | 2 | 3 | 3 | 6 |
|  | every 2d | 3 | 23 | 267 | 2 | 269 | 0 | 2 | 2 | 3 | 5 |
|  | every 1d | 2 | 16 | 133 | 2 | 135 | 0 | 2 | 1 | 3 | 4 |
| $V - S_P$ | every 14d | 12 | 30 | 1,012 | 12 | 1,024 | 1 | 4 | 6 | 3 | 9 |
|  | every 7d | 10 | 31 | 871 | 9 | 880 | 0 | 4 | 5 | 3 | 8 |
|  | every 3d | 7 | 31 | 627 | 6 | 633 | 0 | 3 | 3 | 3 | 6 |
|  | every 2d | 6 | 37 | 508 | 5 | 513 | 0 | 2 | 3 | 3 | 6 |
|  | every 1d | 5 | 46 | 334 | 3 | 337 | 0 | 2 | 2 | 3 | 5 |
| $V - \bar{S}$ | N/A | 15 | 26 | 1,185 | 15 | 1,200 | 1 | 5 | 7 | 3 | 10 |
| **30 vaccines/day** |  |  |  |  |  |  |  |  |  |  |  |
| $V - S_F$ | every 14d | 13 | 31 | 1,307 | 16 | 1,323 | 2 | 6 | 16 | 7 | 23 |
|  | every 7d | 10 | 31 | 1,067 | 11 | 1,078 | 2 | 5 | 13 | 7 | 20 |
|  | every 3 d | 5 | 24 | 630 | 5 | 635 | 2 | 3 | 8 | 7 | 15 |
|  | every 2d | 3 | 23 | 440 | 3 | 443 | 2 | 3 | 6 | 7 | 13 |
|  | every 1d | 2 | 16 | 222 | 2 | 224 | 1 | 2 | 4 | 7 | 11 |
| $V - S_P$ | every 14d | 14 | 31 | 1,381 | 18 | 1,399 | 2 | 6 | 17 | 7 | 24 |
|  | every 7d | 11 | 31 | 1,224 | 13 | 1,237 | 2 | 5 | 14 | 7 | 21 |
|  | every 3d | 6 | 31 | 906 | 8 | 914 | 2 | 4 | 10 | 7 | 17 |
|  | every 2d | 5 | 37 | 729 | 6 | 735 | 1 | 4 | 8 | 7 | 15 |
|  | every 1d | 3 | 44 | 480 | 4 | 484 | 1 | 3 | 5 | 7 | 12 |
| $V - \bar{S}$ | N/A | 17 | 30 | 1,538 | 23 | 1,561 | 2 | 7 | 19 | 8 | 27 |
| **No vaccination** |  |  |  |  |  |  |  |  |  |  |  |
| $\bar{V} - \bar{S}$ | N/A | 20 | 31 | 1,802 | 32 | 1,834 | 2 | 8 | 23 | 9 | 32 |
| $\bar{V} - S_F$ | every 14d | 15 | 31 | 1,622 | 22 | 1,644 | 2 | 7 | 20 | 8 | 28 |
|  | every 7d | 11 | 31 | 1,392 | 16 | 1,408 | 2 | 6 | 16 | 8 | 24 |
|  | every 3d | 6 | 31 | 857 | 8 | 865 | 2 | 4 | 10 | 7 | 17 |
|  | every 2d | 4 | 30 | 594 | 5 | 599 | 2 | 3 | 7 | 7 | 14 |
|  | every 1d | 2 | 23 | 292 | 2 | 294 | 1 | 2 | 4 | 7 | 11 |

Regarding compliance, at only 50% compliance for both vaccination and screening, 60 vaccines/day & weekly screening of the unvaccinated population results in 1,226 infections and a peak of 6 hospitalizations (see Table D in S1 Appendix). Increasing the screening (vaccination) compliance to 95%, while keeping vaccination (screening) compliance at 50%, results in 1,051 (765) infections and 5 (3) peak hospitalizations. Thus, increasing the vaccination compliance reduces the number of infections and the peak number of hospitalizations more substantially than increasing screening compliance, that is, compliance for vaccination and screening do not have symmetric effects.

As expected, total and peak hospitalizations and the total number of infections may increase (decrease) substantially when we consider the worst-case (best-case) scenario; for example, for 60 vaccines/day & 14-day full screening with compliance rates of 75%, the number of infections is 438, 924 and 1,429, and the peak number of hospitalizations is 2, 4 and 7 for the best-, base- and worst-case scenarios, respectively, Table E and Fig B in S1 Appendix.

Fig 1 displays the peak number of hospitalizations versus the number of infections in the base-case scenario, under 60% initial vaccination coverage and 75% screening and vaccination compliance; such analyses can assist decision-makers in selecting an effective strategy based on what is feasible, acceptable, and/or ideal for their college’s setting.

**Fig 1.**
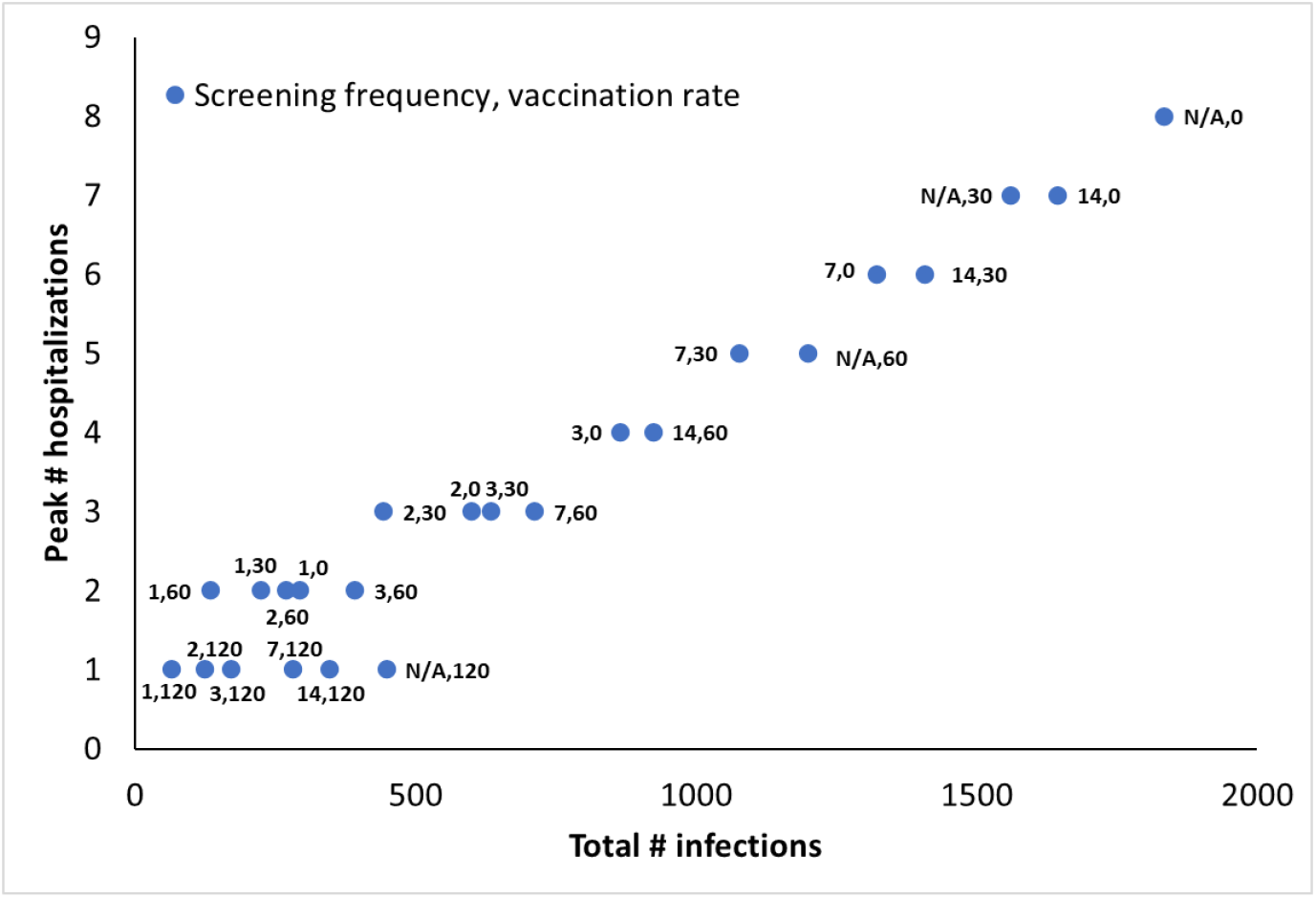
Peak number of hospitalizations versus total number of infections for an initial campus-wide vaccination coverage of 60% (*L*_*s*_ = *L*_*f*_ = 60%) under 75% compliance (*η* = *α* = 75%) for the base-case with full screening

### Impact of Initial Campus-wide Immunity on Strategy Effectiveness

The effectiveness of any strategy is impacted by initial vaccination coverage. For example, at 60% initial vaccine coverage, 120 vaccines/day & no screening lowers the number of infections compared to 60 vaccines/day & weekly screening (Fig 1), however, the opposite is true for 30% initial vaccination coverage (Table C, Table B, Fig B and Fig C in S1 Appendix).

To understand how initial vaccine coverage affects the number of infections, consider, for example, 60 vaccines/day & full weekly screening (strategy *V − S*_*F*_), which results in 2,749, 711, and 13 infections, and 12, 3, and 0 peak hospitalizations in the base-case, for initial vaccination coverage of 30%, 60%, 90%, respectively (Table 4). For low initial vaccination coverage of 30%, augmenting the 60 vaccines/day with aggressive (daily and full) screening still leads to 487 infections over the semester, representing 8.9% of the college’s population, compared to the no-intervention case with 3,512 infections. At 30% initial vaccination coverage, even more aggressive on-campus vaccination (120 vaccines/day) and screening efforts continue to result in a high number of infections: 1,024 to 2,040 for daily partial screening; 275 to 895 with daily full screening; and 1,532 to 1,773 infections with 2-day partial screening in the base- and worst-case scenarios, respectively considering reproduction numbers of 4.75 to 6.75.

**Table 4.** Total number of infections and peak number of hospitalizations for the 80-day semester for some strategies, implemented at various screening frequencies and a vaccination rate of 60 vaccines/day, under the base-case scenario, considering 75% compliance (*η* = *α* = 75%) and initial campus-wide immunity (*L*_*s*_ = *L*_*f*_) of 30%, 60%, 90%.

| | | $L_s : 30\% , L_f : 30\%$ | | $L_s : 60\% , L_f : 60\%$ | | $L_s : 90\% , L_f : 90\%$ | |
| --- | --- | --- | --- | --- | --- | --- | --- |
| Strategy | Test Frequency | Total number of infections | Peak number of hospitalizations | Total number of infections | Peak number of hospitalizations | Total number of infections | Peak number of hospitalizations |
| 60 vaccines/day |  |  |  |  |  |  |  |
| $V - S_F$ | every 14d | 3,014 | 13 | 924 | 4 | 15 | 0 |
|  | every 7d | 2,749 | 12 | 711 | 3 | 13 | 0 |
|  | every 3 d | 1,822 | 8 | 392 | 2 | 10 | 0 |
|  | every 2d | 1,185 | 5 | 269 | 2 | 8 | 0 |
|  | every 1d | 487 | 3 | 135 | 2 | 5 | 0 |
| $V - S_P$ | every 14d | 3,075 | 14 | 1,024 | 4 | 15 | 0 |
|  | every 7d | 2,938 | 13 | 880 | 4 | 14 | 0 |
|  | every 3d | 2,536 | 10 | 633 | 3 | 12 | 0 |
|  | every 2d | 2,219 | 9 | 513 | 2 | 10 | 0 |
|  | every 1d | 1,603 | 6 | 337 | 2 | 7 | 0 |
| $V - \overline{S}$ | N/A | 3,184 | 15 | 1200 | 5 | 17 | 0 |

## Discussion

Integrating routine screening with vaccination during various phases of a pandemic can play a key role in managing the spread of the infection and mitigating the impact of the pandemic. Results from the extended compartmental model in this study suggest that routine screening and vaccination work synergistically to contain a SARS-CoV-2 outbreak and achieve a level of success that neither strategy can achieve alone. However, to maximize the effectiveness, the strategy needs to be customized for each setting, considering community and disease characteristics.

Vaccination is an effective and sustainable tool for fighting disease outbreaks [60], and reduces infection transmission [15, 57] and the consequent hospitalization and death [18, 28, 31]. Although it is of critical importance to deliver and administer the available vaccines in a swift manner, vaccination efforts might be limited by resource scarcity [4, 41, 44], especially during the earlier stages of a pandemic, as well as vaccine hesitancy due, for example, to lack of trust and confidence in vaccines or health systems, or fear of side effects [9, 38, 42, 55]. Currently, 60% of the U.S. population is fully vaccinated for COVID-19, with 71% of the population receiving at least one dose of a two-dose vaccine [54], and only 17% receiving a booster shot [32], despite that vaccines have been available for every adult in the U.S. since April 2021 [2], and for 12 and over since May 2021 [27]. Because the proportion of the fully vaccinated population is still below the 70% *−* 85% threshold estimated to contain an infection [21, 25], and because vaccines do not provide immediate protection against the infection [7] and vaccine-induced immunity wanes over time [20], it is critical to understand how vaccination, screening, and isolation can be utilized in the containment of a SARS-CoV-2 outbreak for a college campus, and whether such on-campus efforts can substitute for low vaccination rates at the start of the semester.

Our results suggest that the effectiveness of any intervention strategy, relatedly the college’s ability to offer a viable in-person teaching experience, depends heavily on initial vaccination coverage. For the study population of 5,455 over an 80-day semester, with low initial vaccination coverage (30% in our study), even aggressive on-campus vaccination and screening efforts with moderate compliance (75%) result in a high number of infections: 1,024 to 2,040 for routine daily screening of the unvaccinated population; 275 to 895 with extended screening to also cover the newly vaccinated population; and 1,532 to 1,773 infections for every 2-day screening of the unvaccinated population, in the base- and worst-case scenarios (see Table F in S1 Appendix), respectively considering reproduction numbers of 4.75 and 6.75, representative of the COVID-19 Delta variant [24]. These results suggest that aggressive on-campus interventions throughout the semester cannot typically compensate for low initial vaccination rates, that is, when starting the semester with a highly susceptible population, especially when the disease reproduction number is relatively high. Further, with the emergence of the COVID-19 Omicron variant, it is likely that the reproduction number will increase and/or the effective vaccine coverage will decrease if a booster shot is needed to maximize vaccine efficacy. These findings underscore the importance of heavily promoting, even mandating, full vaccination at the start of the semester.

Our study also highlights the need to customize on-campus vaccination and screening efforts based on community and disease characteristics. For example, at a moderate initial vaccine coverage of 60%, an integrated vaccination and screening strategy of 60 vaccines per day and 2-day screening dominates both an aggressive vaccination-only (at 120 vaccines per day) strategy and an aggressive screening-only (daily screening) strategy in terms of the number of infections. Of course, the dominating strategy depends on initial vaccine coverage, and needs to be carefully chosen. Equally important is the finding that integrating routine screening (both partial and full, at every 14 days or less) with daily vaccination is beneficial at all vaccination rates and initial vaccine coverage levels, and is associated with a lower number of infections and improved peak number of hospitalization over an 80-day semester. This result suggests that even a mild regime of routine screening, followed by immediate isolation of the positive cases, can complement vaccination and help mitigate the spread of the disease and alleviate the burden on the hospital systems, especially in institutions like college campuses, nursing homes, prisons, or military bases in which higher contact rates are expected.

Another important policy question regarding the addition of routine screening to vaccination is whether the vaccinated population should continue to be screened until they build immunity against the infection. The findings in this study suggest that including the vaccinated population in the screening pool can potentially decrease the total number of infections, although the degree of these benefits varies with screening frequency as well as the daily number of vaccines administered. Another policy decision related to an integrated intervention effort is the frequency of administering each intervention. Although the daily vaccination rate is expected to be constrained by the number of available doses as well as the public’s willingness to be vaccinated, especially in the early stages of vaccination efforts, screening resources are generally abundant, and their frequency may be set considering the health return and feasibility of each strategy. The results of this study show that the incremental benefit of more frequent screening diminishes as the daily vaccination rate increases.

Most colleges have established plans and guidelines for containing a campus-wide outbreak of SARS-CoV-2. While many colleges mandate either vaccination, or medium to high frequency screening if unvaccinated [30, 49], others require neither, even in regions with low vaccination rates. These differences underscore the importance of an outcome-based analysis of different potential interventions, such as the study conducted in this paper, to guide the decision-maker (e.g., college) in the most appropriate strategy given their unique characteristics. Our findings imply that multiple factors and constraints need to be included in assessing the best strategy and measures for fighting disease outbreaks. These factors may include the daily or total number of infections a college is willing to tolerate, the peak level of hospitalization that local hospitals can cope with, screening and vaccination capacity that the campus is able to achieve, initial vaccine coverage on campus, and the transmission severity scenario. It should also be considered that a partially-vaccinated campus environment (i.e., without a vaccine mandate) could provide a false sense of security for the overall student population, including those who are unvaccinated, and, as a result, infections may rise quite rapidly during the academic semester. Our results also suggest that increasing vaccination compliance has a more substantial effect in reducing the number of infections than increasing routine screening compliance. This implies that compliance for vaccination and screening do not have symmetric effects.

Finally, early studies indicate that existing COVID-19 vaccines, without a booster shot, may have low efficacy for the emerging Omicron variant [12, 51]. Our findings indicate that a sufficiently high level of initial immunity, through full vaccination (which may imply a booster shot), before arriving on campus, is key to reducing the spread of a disease on a college campus. For low initial vaccine coverage (e.g., 30% in our study), even aggressive on-campus vaccination and screening efforts may not be sufficient to keep the spread under control.

Recently, several U.S. colleges have added the booster shot to their current mandate of full vaccination for COVID-19, thus requiring all students and faculty/staff to take the booster shot prior to the start of the Spring 2021/2022 semester, e.g., [5, 16], whereas others have delayed in-person teaching until a safe re-opening is feasible, e.g., [53]. Given the uncertain and dynamic nature of the pandemic, our analysis indeed confirms that academic institutions need to remain flexible, and adjust their plans as needed prior to the start of each academic semester, and take immediate action based on the most recent guidelines and the current state of the pandemic. If done right, an integrated routine screening and vaccination strategy can contain any potential disease outbreak and maintain a safe environment for the local community.

## Limitations

We use a compartmental model, which, by definition, is based on assumptions such as homogeneous mixing, within and between, student and faculty compartments, and uniform contact, transmission, hospitalization, and fatality rates for all individuals within a compartment. We also assume that the vaccine-induced immunity follows a step function: zero immunity in the earlier stages after vaccination, followed by full immunity after two doses (if the vaccine is effective for the individual), which takes a stochastic amount of time. We do not consider waning immunity or booster shots. While we model noncompliance with on-campus screening and vaccination guidelines, we assume that the population with symptoms or positive test results fully comply with isolation recommendations. We do not consider other mitigation tools, such as mask wearing and contact tracing.

While many parameters in our model are assumed deterministic, sensitivity analysis has been conducted on key parameters, which shows that effective strategies are robust to a wide range of parameter values. Therefore, we expect the qualitative results to continue to hold under stochasticity in parameter values. Still it would be interesting to study our strategies by explicitly modeling the stochasticity.

Finally, we consider only two groups (students and faculty), as this captures the main heterogeneity that is present on a college campus. However, more groups (compartments) can be added to the model, but this comes at the expense of increased complexity and data requirements. It would be interesting to develop a more detailed model through, for example, an agent-based simulation, which is particularly useful for modeling this type of complexity [39].

## Conclusions

Integrated routine screening and vaccination is effective for containing a SARS-CoV-2 outbreak during an academic semester on a college campus, as long as initial vaccination coverage is sufficiently high, depending on the reproduction rate of the disease. Vaccination and routine screening work synergistically to achieve a level of success that neither strategy can achieve alone. However, the incremental benefit of integrated screening decreases as vaccination rate and/or initial vaccination coverage increases. Further, to maximize efficacy, the intervention strategy needs to be customized for the setting, considering community and disease characteristics, including the vaccine coverage at the start of the semester, estimated screening and vaccination compliance rates, as well as the threshold number of infections and the peak number of hospitalization a campus is willing to tolerate. The model developed in this paper can be used to establish effective strategies for containing disease outbreaks in high-density settings, including college campuses, nursing homes, and military bases.

## Data Availability

All data produced in the present work are contained in the manuscript and the appendix

## Supporting Information

**S1 Appendix. Supplementary Online Content**

## Notes

### Competing Interest Statement

The authors have declared no competing interest.

### Funding Statement

This study did not receive any funding

