## Supplementary material for "Benefits of integrated screening and vaccination for infection control": S1 Appendix

### S1 Appendix: Supplementary Online Content

#### Model Description

We developed an extended SEIR (Susceptible, Exposed, Infectious, Removed) framework to model the COVID-19 spread in a population composed of two homogeneous groups (faculty and students) under protective and preventative interventions including screening, isolation, and vaccination. The model tracks the population of at-risk individuals while they transition through different health states, or *compartments*, where each transition is modeled probabilistically and the overall flow is governed by a series of difference equations. Our model provides an extension to the conventional SIR frameworks with mass vaccination by generalizing the SEIR compartmental model of [1], which was introduced to study the epidemiology and natural history of COVID-19 infection. In particular, we expanded the compartmental model in [1] in the following ways, which are also illustrated in the flow chart given in Fig A:

- Inclusion of vaccination performed concurrently with isolation and screening.
- Modeling the probabilistic time after vaccination for the body to build protection (immunity).
- Distinguishing between vaccinated (but not fully immune) and unvaccinated individuals in the active transmission and screening pool.
- Recognizing the potential "immunity unknowing" by differentiating between the individuals with a detected infection (through either having symptoms or having a positive

PCR test) or vaccination and the ones that got recovered from the infection unknowingly because of the absence of symptoms or a positive PCR test.

- Inclusion of the option for vaccinated individuals to be tested.
- Improved accuracy of the model and the equations depicting the model.
- Recognizing the population-based disparities in the rates of disease spread, hospitalization, and mortality by considering two groups of population, i.e., students and faculty, and their interactions.
- Modeling the vaccine and screening compliance.
- Modeling the imperfect vaccine efficacy.
- Inclusion of the option for hospitalization of symptomatic cases.

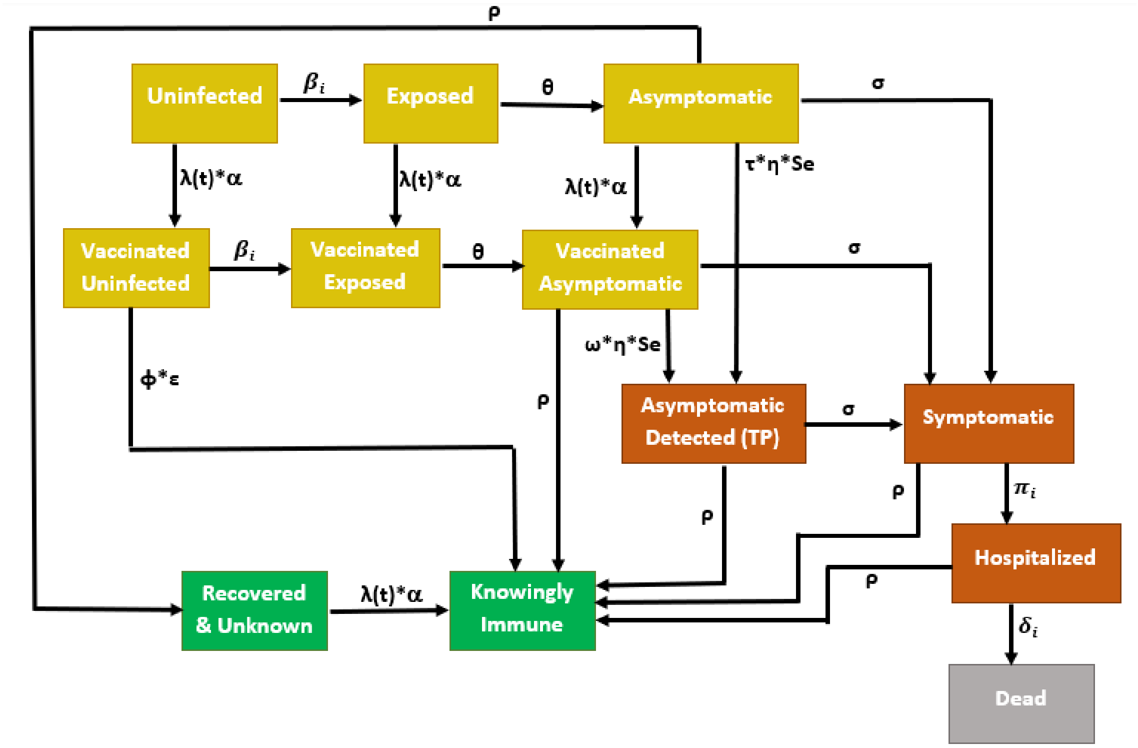


**Fig A:** Flow Diagram of the Extended SEIR Model

**Compartments**

We defined a total of 15 model compartments for each population subgroup i, where $i \in \{students, faculty\}$, and kept the inherited compartments from [1] with their original names to underline the similarities and novelties in between the two models.

**Unvaccinated transmission and screening pool.** All individuals in this pool can be vaccinated. This pool can also be tested for infection.

- $U_i$: Uninfected, unvaccinated, susceptible individuals
- $E_i$: Exposed, unvaccinated, asymptomatic, non-infectious
- $A_i$: Asymptomatic, unvaccinated, infected

**Vaccinated transmission and screening pool.** All individuals in this pool are assumed to be vaccinated but not fully immune yet. Depending on the screening strategy, these individuals can be tested for infection. Together with the *Unvaccinated transmission and screening pool*, these pools involve all individuals who can transmit infections or get infected.

- $VU_i$: Vaccinated, uninfected, susceptible individuals.
- $VE_i$: Vaccinated, exposed, asymptomatic, non-infectious.
- $VA_i$: Vaccinated, asymptomatic, infected.

**Unknowingly recovered screening pool.** All the individuals in this pool are assumed to be recovered from infection unknowingly. Together with *Unvaccinated transmission and screening pool* and *Vaccinated transmission and screening pool*, these pools are assumed to include all individuals that can be tested for infection.

- $RU_i$: Recovered & Unknown. Individuals in this compartment were unknowingly infected and recovered.

**Isolation pool.** All individuals in this pool are assumed to be isolated from each other as well as from the other pools. No transmission takes places within this pool.

- $FP_i$: False-Positive result, uninfected.
- $FPVU_i$: False-Positive, Vaccinated & Uninfected.
- $FPRU_i$: False-Positive, Recovered & Unknown.
- $TP_i$ (Asymptomatic Detected): True Positive result, asymptomatic, infected.
- $S_i$: Symptomatic, detected.
- $H_i$: Hospitalized.

**Removed pool.** Individuals in this pool are assumed to be immune for at least 80 days (duration of the semester). Accordingly, we assume that no transmission, screening or vaccination (including any boosters) takes place among the individuals in this pool.

- $D_i$: Dead.
- $KI_i$: Knowingly immune. Individuals in this compartment were either knowingly infected and recovered or vaccinated and built immunity.

A summary of the compartmental flow assumptions for each compartment is given in Table A.

**Table A:** Summary of Compartmental Flow Assumptions

| **Compartment** | **In screening pool?** | **In vaccination pool?** | **In isolation pool?** |
|---|---|---|---|
| ($U_i$)–Uninfected | Yes | Yes | No |
| ($E_i$)–Exposed | Yes | Yes | No |
| ($A_i$)–Asymptomatic | Yes | Yes | No |
| ($RU_i$)–Recovered & Unknown | Yes | Yes | No |
| ($VU_i$)–Vaccinated Uninfected | Depends | No | No |
| ($VA_i$)–Vaccinated Asymptomatic | Depends | No | No |
| ($VE_i$)–Vaccinated Exposed | Depends | No | No |
| ($FP_i$)–False-Positive | No | No | Yes |
| ($FPVU_i$)–False-Positive, Vaccinated & Uninfected | No | No | Yes |
| ($FPRU_i$)–False-Positive, Recovered & Unknown | No | No | Yes |
| ($TP_i$)–True-Positive, Asymptomatic Detected | No | No | Yes |
| ($S_i$)–Symptomatic | No | No | Yes |
| ($H_i$)–Hospitalized | No | No | Yes |
| ($KI_i$)–Knowingly Immune | No | No | No |
| ($D_i$)–Dead | No | No | No |

**Model Parameters**

Subscript $i, j \in \{\text{students}, \text{faculty}\}$ denotes the subgroup of the population. Parameters without these subscripts are the same for both subgroups.

$\beta_{j,i}$: rate at which infected subjects in group $j$ contact subjects from group $i$ and infect them

$\tau$: rate at which unvaccinated subjects in the screening pool are screened for infection

$\theta$: rate at which exposed subjects advance to the Asymptomatic & infectious compartment

$\delta_i$: rate at which subjects of group $i$ in the Hospitalized compartment die

$\pi_i$: rate at which subjects of group $i$ in the Symptomatic compartment get hospitalized

$\rho$: rate at which infected subjects recover from disease and are removed

$\sigma$: rate of symptom onset for infected subjects

$\mu$: rate at which false-positives are returned to the Uninfected compartment

$sens$: sensitivity of the screening test

$spec$: specificity of the screening test

I(t): an indicator function which assumes a value 1 if an exogenous shock takes place in cycle t; 0 Otherwise

$X$: number of imported infections to group $i$ in a given exogenous shock

$\omega$: rate at which vaccinated subjects in the screening pool are screened for infection

$\lambda(t)$: rate at which eligible subjects are vaccinated

$\phi$: rate at which vaccinated subjects advance to the "Immune & known" compartment,

$\alpha$: vaccine compliance rate

$\eta$: screening compliance rate

$\epsilon$: vaccine efficacy

The model uses a cycle time of 8 hours. Screening rates $\tau$ and $\omega$ are assumed to be static, whereas the vaccination rate $\lambda(t)$ is time-dependent so that a constant number of vaccines are administered per day. Certain parameter values are varied in the analysis to simulate different strategies or scenarios, e.g., vaccination rate, screening frequency, etc.

##### Governing Equations

The following defines the governing equations for the model depicted in Fig A, where $i, j \in \{\text{students}, \text{faculty}\}$ and $Z_i(t) := U_i(t) + VU_i(t) + E_i(t) + VE_i(t) + A_i(t) + VA_i(t) + RU_i(t) + KI_i(t)$.

$$
\begin{aligned}
U_i(t+1) &= U_i(t) \cdot \left[1 - \sum_j \left[\beta_{j,i} \cdot \frac{A_j(t) + VA_j(t)}{Z_j(t)}\right] - \lambda(t) \cdot \alpha\right] \\
&\quad - U_i(t-1) \cdot \tau \cdot \eta \cdot (1 - spec) + \mu \cdot FP_i(t) - X \cdot I(t+1) \\
E_i(t+1) &= E_i(t) \cdot [1 - \theta - \lambda(t) \cdot \alpha] + \sum_j \left[\beta_{j,i} \cdot \frac{U_i(t) \cdot [A_j(t) + VA_j(t)]}{Z_j(t)}\right] + X \cdot I(t+1) \\
A_i(t+1) &= A_i(t) \cdot [1 - \sigma - \rho - \lambda(t) \cdot \alpha] - A_i(t-1) \cdot \tau \cdot \eta \cdot sens + E_i(t) \cdot \theta \\
FP_i(t+1) &= FP_i(t) \cdot [1 - \mu] + U_i(t-1) \cdot \tau \cdot \eta \cdot (1 - spec) \\
TP_i(t+1) &= TP_i(t) \cdot [1 - \sigma - \rho] + A_i(t-1) \cdot \tau \cdot \eta \cdot sens + VA_i(t-1) \cdot \omega \cdot \eta \cdot sens \\
S_i(t+1) &= S_i(t) \cdot [1 - \rho - \pi_i] + \sigma \cdot [TP_i(t) + A_i(t) + VA_i(t)] \\
H_i(t+1) &= H_i(t) \cdot [1 - \rho - \delta_i] + \pi_i \cdot S_i(t) \\
KI_i(t+1) &= KI_i(t) + \rho \cdot [TP_i(t) + S_i(t) + H_i(t) + VA_i(t)] + \lambda(t) \cdot \alpha \cdot RU_i(t) + \phi \cdot \epsilon \cdot VU_i(t) \\
RU_i(t+1) &= RU_i(t) \cdot (1 - \lambda(t) \cdot \alpha) + \rho \cdot A_i(t) - RU_i(t-1) \cdot \tau \cdot \eta \cdot (1 - spec) + \mu \cdot FPRU_i(t) \\
D_i(t+1) &= D_i(t) + \delta_i \cdot H_i(t) \\
FPVU_i(t+1) &= FPVU_i(t) \cdot [1 - \mu] + VU_i(t-1) \cdot \omega \cdot \eta \cdot (1 - spec) \\
FPRU_i(t+1) &= FPRU_i(t) \cdot [1 - \mu] + RU_i(t-1) \cdot \tau \cdot \eta \cdot (1 - spec) \\
VU_i(t+1) &= VU_i(t) \cdot \left[1 - \phi \cdot \epsilon - \sum_j \left[\beta_{j,i} \cdot \frac{VA_j(t) + A_j(t)}{Z_j(t)}\right]\right] \\
&\quad + \lambda(t) \cdot \alpha \cdot U_i(t) - VU_i(t-1) \cdot \omega \cdot \eta \cdot (1 - spec) + \mu \cdot FPVU_i(t) \\
VE_i(t+1) &= VE_i(t) \cdot (1 - \theta) + \sum_j \left[\beta_{j,i} \cdot \frac{VU_i(t)[VA_j(t) + A_j(t)]}{Z_j(t)}\right] + \lambda(t) \cdot \alpha \cdot E_i(t)
\end{aligned}
$$

$$VA_i(t+1) = VA_i(t)\cdot(1-\sigma-\rho) + \lambda(t)\cdot\alpha\cdot A_i(t) - VA_i(t-1)\cdot\omega\cdot\eta\cdot sens + VE_i(t)\cdot\theta$$

$$N = \sum_i \Big[U_i + E_i + A_i + S_i + TP_i + FP_i + KI_i + H_i + RU_i + D_i + FPVU_i + FPRU_i + VU_i + VA_i + VE_i\Big]$$

**Initial Conditions**

- $KI_i(0) = \begin{cases} \{1500, 3000, 4500\}, & i = students \\ \{136, 273, 409\}, & i = faculty \end{cases}$
- $A_i(0) = \begin{cases} 10, & i = students \\ 1, & i = faculty \end{cases}$
- $U_i(0) = \begin{cases} 5000 - A_i(0) - KI_i(0), & i = students \\ 455 - A_i(0) - KI_i(0), & i = faculty \end{cases}$

All other compartments are initially empty. Accordingly, $N = 5000 + 455 = 5455$.

**Estimating Key Rate Parameters**

The following parameters are inherited from [1],

- $\sigma$, rate of symptom onset for infected individuals: $\sigma$ is estimated by solving $\sigma/(\sigma+\rho) = 30\%$, where 30% [2] is the probability of getting symptoms after an infection and $\rho$ is the rate of recovery. Since the time to recovery is assumed to be 14 days and the day is composed of 3 eight hours cycles, we have $\rho = 1/(3\cdot 14)$, which gives $\sigma = 0.0102$.
- $\tau$, rate at which unvaccinated individuals in the screening pool are screened for infection: It is given by $\tau = \frac{1}{3\cdot f}$, where f is the screening frequency, which can be *daily, every 2 days, every 3 days, every 7 days or every 14 days*

In addition, we use the following estimates.

- $\beta_{j,i}$, rate at which infected subjects in group $j$ contact subjects from group $i$ and infect them, where $i, j \in \{\text{students}, \text{faculty}\}$. It varies based on the gravity of transmission (best, base or worst case scenario) which is represented in terms of reproduction number $R_{T(j,i)}$. For notational simplicity, we denote students with 's' and faculty with 'f' in the remainder of the paragraph. Based on the type of dynamics, we assume that $R_{T(s,s)} = 5.4$, $R_{T(s,f)} = 0.6$, $R_{T(f,f)} = 0.32$ and $R_{T(f,s)} = 2.88$. Thus, we get that the overall $R_{T(s)} = 6$ and $R_{T(f)} = 3.2$ assuming a 11:1 student to faculty ratio (based on [3]). As a result, the population reproduction number $R_T$ for the base-case is $R_T = 0.09 \cdot R_{T(f)} + 0.91 \cdot R_{T(s)} = 5.75$. $\beta_{j,i}$ is estimated by solving $R_{T(j,i)} = \beta_{j,i}/(\sigma + \rho)$, which gives $\beta_{s,s} = 0.184$, $\beta_{s,f} = 0.0204$, $\beta_{f,f} = 0.0109$ and $\beta_{f,s} = 0.098$ for the base-case scenario.

  For the best-case and worst-case scenarios, we set $R_T$ to 6.75 and 4.75, respectively. Accordingly, under these scenarios, $R_{T(s)}$ becomes 5 and 7 and $R_{T(f)}$ becomes 2.2 and 4.2, respectively. Therefore, we have $R_{T(s,s)} = 4.5$, $R_{T(s,f)} = 0.5$, $R_{T(f,f)} = 0.22$ and $R_{T(f,s)} = 1.98$ (i.e., $\beta_{s,s} = 0.153$, $\beta_{s,f} = 0.017$, $\beta_{f,f} = 0.00748$ and $\beta_{f,s} = 0.0673$) for the best-case scenario and $R_{T(s,s)} = 6.3$, $R_{T(s,f)} = 0.7$, $R_{T(f,f)} = 0.42$ and $R_{T(f,s)} = 3.78$ (i.e., $\beta_{s,s} = 0.214$, $\beta_{s,f} = 0.0238$, $\beta_{f,f} = 0.0143$ and $\beta_{f,s} = 0.128$) for the worst-case scenario.

- $\pi_i$, rate at which subjects in the *Symptomatic* compartment of group $i$ die, where $i \in \{students, faculty\}$. These rates are calculated based on the hospitalization rates of 1.4% and 8.4% for students and faculty, respectively [4, 5]. The hospitalization rates are obtained by the current and cumulative rate of COVID-19 hospitalizations per age group (with respect to the total population) [4] and the total percentage of infections per age group [5]. For each age group, the hospitalization rate (among the infected subjects) is the ratio of the corresponding rate of hospitalization to the percentage of infections. Assuming all university students to be in the 18-29 age group, we get a hospitalization rate of 1.4% for students. For the faculty, we consider that their age range covers multiple age groups (30-39, 40-49, 50-64, 65-74 and 75-84) [6]. The percentage of faculty in each of these five age groups are estimated as 17%, 33%, 37%, 10%, and 3%, respectively, with the corresponding hospitalization rates of 2.9%, 4.8%, 5.7%, 21.5% and 69.3%, respectively. Using a weighted average yields a faculty hospitalization rate of 8.4%. For $i \in \{\text{students}, \text{faculty}\}$, $\pi_i$ is the solution to $[\sigma/(\rho+\sigma)] \cdot [\pi_i/(\rho+\pi_i)] = HospitalizationRate_i$, which gives $\pi_{student} = 0.001166$ and $\pi_{faculty} = 0.009263$.

- $\delta_i$, rate at which subjects in the *Hospitalized* compartment of group $i$ die. It is based on the fatality rate of 0.05% and 2% for students and faculty respectively [7,8]. The fatality rates are obtained using the current number of COVID-19 deaths per age group [7] and the current number of infections per age group [8]. For each age group, the fatality rate is the ratio of the corresponding number of deaths to the number of infections. Assuming all university students to be in the 18-29 age group, we get a fatality rate of 0.05% for students. Similar to the calculation of $\pi_i$, we consider five different age groups for the faculty with the corresponding fatality rates of 0.2%, 0.6%, 2%, 6%, and 14%, respectively. Using a weighted average yields a faculty fatality rate of 2%. For $i \in \{\text{students}, \text{faculty}\}$, $\delta_i$ is the solution to $[\sigma/(\rho+\sigma)] \cdot [\delta_i/(\rho+\delta_i)] \cdot [\pi_i/(\rho+\pi_i)] = FatalityRate_i$, which gives $\delta_{students} = 0.0008817$ and $\delta_{faculty} = 0.00744$.

- $\epsilon$, vaccine efficacy: We only consider 2-dose vaccines (Pfizer and Moderna) because of the scarce application ($< 4\%$) of 1-dose vaccine (Janssen) compared to the other two (58% and 38%, respectively) in the US [9]. Accordingly, the vaccine efficacy $\epsilon$ is calculated as the 60%-40% weighted average of Pfizer's efficacy of 95% [10] and Moderna's of 94.1% [11]. Therefore $\epsilon = 0.6 \cdot 95\% + 0.4 \cdot 94.1\% = 94.64\%$.

- $\eta$, compliance rate of screening: We use a baseline value of $\eta = 0.75$, and perform a sensitivity analysis at several levels of screening compliance using $\eta = \{0.5, 0.75, 1\}$.

- $\alpha$, compliance rate of vaccination: We use a baseline value of $\alpha = 0.75$, and perform a sensitivity analysis at several levels of vaccination compliance using $\alpha = \{0.5, 0.75, 1\}$.

- $\phi$, average time after vaccination for the body to build full immunity: It is estimated to take 5 or 6 weeks on average to gain full immunity after being vaccinated with Pfizer or Moderna, respectively [12]. Accordingly, using a weighted average, we obtain $\phi = \frac{1}{3*7*(0.6*5+0.4*6)} = 0.0089$.

- $\omega$, rate at which vaccinated individuals in the screening pool are screened for infection: $\omega = \tau$ when vaccinated individuals are tested, $\omega = 0$ otherwise.

- $\lambda(t)$, rate at which eligible individuals are vaccinated: $\lambda(t)$ is time-dependant and updated dynamically such that the number of vaccines given per time period (8 hours) is fixed. Initially, we set $\lambda(t+1) = \frac{VPP}{\sum_i [U_i(t)+E_i(t)+A_i(t)+RU_i(t)]}$ where $VPP$ represents the number of vaccines per time period. In order to be able to vaccinate most of the students by the end of the semester, we need to administer at least 60 vaccines per day. Accordingly, $VPP = \frac{60}{3} = 20$ is used in the base model of our study, and a sensitivity analysis is performed over this value.

Fig A presents a flow diagram of the extended SEIR model. For clarity, Fig A does not include the false-positive compartments. Furthermore, for the compartments separately defined for students and faculty, only one compartment is shown in the figure.

**Table B:** Total number of infections and peak number of hospitalizations for the 80-day semester for all strategies, implemented at various screening frequencies, vaccination rates and initial student vaccination coverage ($L_s$), under the base-case scenario, considering 75% compliance ($\eta = \alpha = 75\%$) and initial faculty immunity ($L_f$) of 30%.

| | | $L_s: 30\%, L_f: 30\%$ | | $L_s: 60\%, L_f: 30\%$ | | $L_s: 90\%, L_f: 30\%$ | |
|---|---|---|---|---|---|---|---|
| Strategy | Test Frequency | Total number of infections | Peak number of hospitalizations | Total number of infections | Peak number of hospitalizations | Total number of infections | Peak number of hospitalizations |
| **120 vaccines/day** | | | | | | | |
| $V - S_F$ | every 14d | 2,419 | 7 | 403 | 1 | 9 | 0 |
| | every 7d | 2,019 | 6 | 324 | 1 | 8 | 0 |
| | every 3 d | 1,121 | 4 | 197 | 1 | 6 | 0 |
| | every 2d | 695 | 3 | 141 | 1 | 5 | 0 |
| | every 1d | 275 | 2 | 74 | 1 | 3 | 0 |
| $V - S_P$ | every 14d | 2,564 | 8 | 445 | 1 | 9 | 0 |
| | every 7d | 2,348 | 7 | 391 | 1 | 8 | 0 |
| | every 3d | 1,844 | 5 | 296 | 1 | 7 | 0 |
| | every 2d | 1,532 | 4 | 238 | 1 | 6 | 0 |
| | every 1d | 1,024 | 3 | 163 | 1 | 4 | 0 |
| $V - \overline{S}$ | N/A | 2,773 | 9 | 525 | 1 | 9 | 0 |
| **60 vaccines/day** | | | | | | | |
| $V - S_F$ | every 14d | 3,014 | 13 | 1,026 | 5 | 33 | 0 |
| | every 7d | 2,749 | 12 | 793 | 4 | 29 | 0 |
| | every 3 d | 1,822 | 8 | 434 | 3 | 21 | 0 |
| | every 2d | 1185 | 5 | 294 | 2 | 17 | 0 |
| | every 1d | 487 | 3 | 143 | 2 | 9 | 0 |
| $V - S_P$ | every 14d | 3,075 | 14 | 1,126 | 5 | 36 | 0 |
| | every 7d | 2,938 | 13 | 974 | 4 | 33 | 0 |
| | every 3d | 2,536 | 10 | 708 | 3 | 26 | 0 |
| | every 2d | 2,219 | 9 | 570 | 3 | 23 | 0 |
| | every 1d | 1,603 | 6 | 376 | 2 | 16 | 0 |
| $V - \overline{S}$ | N/A | 3,184 | 15 | 1,306 | 6 | 41 | 0 |
| **30 vaccines/day** | | | | | | | |
| $V - S_F$ | every 14d | 3,228 | 14 | 1,412 | 6 | 90 | 1 |
| | every 7d | 3,025 | 13 | 1,162 | 5 | 73 | 1 |
| | every 3 d | 2,195 | 9 | 680 | 3 | 48 | 1 |
| | every 2d | 1,485 | 6 | 470 | 3 | 36 | 1 |
| | every 1d | 599 | 3 | 233 | 2 | 21 | 1 |
| $V - S_P$ | every 14d | 3,255 | 14 | 1,478 | 7 | 97 | 1 |
| | every 7d | 3,126 | 13 | 1,317 | 6 | 85 | 1 |
| | every 3d | 2,683 | 10 | 984 | 4 | 65 | 1 |
| | every 2d | 2,261 | 8 | 791 | 4 | 53 | 1 |
| | every 1d | 1,303 | 5 | 494 | 3 | 36 | 1 |
| $V - \overline{S}$ | N/A | 3,350 | 15 | 1,641 | 8 | 119 | 1 |
| **No vaccination** | | | | | | | |
| $\overline{V} - \overline{S}$ | N/A | 3,512 | 16 | 1,903 | 9 | 318 | 3 |
| $\overline{V} - S_F$ | every 14d | 3,427 | 15 | 1,719 | 7 | 249 | 2 |
| | every 7d | 3,275 | 14 | 1,481 | 6 | 201 | 2 |
| | every 3d | 2,561 | 10 | 909 | 4 | 129 | 2 |
| | every 2d | 1,815 | 7 | 626 | 3 | 98 | 2 |
| | every 1d | 723 | 3 | 303 | 2 | 56 | 2 |

**Table C:** Total number of infections and peak number of hospitalizations for the 80-day semester for all strategies, implemented at various screening frequencies, vaccination rates and initial faculty vaccination coverage ($L_f$), under the base-case scenario, considering 75% compliance ($\eta = \alpha = 75\%$) and initial student immunity ($L_s$) of 30%.

| | | $L_s: 30\%, L_f: 30\%$ | | $L_s: 30\%, L_f: 60\%$ | | $L_s: 30\%, L_f: 90\%$ | |
|---|---|---|---|---|---|---|---|
| Strategy | Test Frequency | Total number of infections | Peak number of hospitalizations | Total number of infections | Peak number of hospitalizations | Total number of infections | Peak number of hospitalizations |
| **120 vaccines/day** | | | | | | | |
| $V - S_F$ | every 14d | 2,419 | 7 | 2,282 | 6 | 2,143 | 6 |
| | every 7d | 2,019 | 6 | 1,879 | 5 | 1,739 | 5 |
| | every 3 d | 1,121 | 4 | 1,032 | 3 | 947 | 3 |
| | every 2d | 695 | 3 | 641 | 3 | 594 | 2 |
| | every 1d | 275 | 2 | 261 | 2 | 244 | 2 |
| $V - S_P$ | every 14d | 2,564 | 8 | 2,423 | 7 | 2,303 | 6 |
| | every 7d | 2,348 | 7 | 2,211 | 6 | 2,068 | 5 |
| | every 3d | 1,844 | 5 | 1,709 | 4 | 1,599 | 4 |
| | every 2d | 1,532 | 4 | 1,414 | 3 | 1,298 | 3 |
| | every 1d | 1,024 | 3 | 926 | 2 | 851 | 2 |
| $V - \overline{S}$ | N/A | 2,773 | 9 | 2,653 | 8 | 2,535 | 7 |
| **60 vaccines/day** | | | | | | | |
| $V - S_F$ | every 14d | 3,014 | 13 | 2,927 | 12 | 2,842 | 12 |
| | every 7d | 2,749 | 12 | 2,647 | 11 | 2,549 | 10 |
| | every 3 d | 1,822 | 8 | 1,718 | 7 | 1,617 | 7 |
| | every 2d | 1,185 | 5 | 1,113 | 5 | 1,042 | 5 |
| | every 1d | 487 | 3 | 463 | 3 | 434 | 3 |
| $V - S_P$ | every 14d | 3,075 | 14 | 2,993 | 13 | 2,915 | 12 |
| | every 7d | 2,938 | 13 | 2,879 | 12 | 2,764 | 11 |
| | every 3d | 2,536 | 10 | 2,431 | 10 | 2,340 | 9 |
| | every 2d | 2,219 | 9 | 2,116 | 8 | 2,028 | 8 |
| | every 1d | 1,603 | 6 | 1,517 | 6 | 1,443 | 5 |
| $V - \overline{S}$ | N/A | 3,184 | 15 | 3,105 | 13 | 3,040 | 13 |
| **30 vaccines/day** | | | | | | | |
| $V - S_F$ | every 14d | 3,228 | 14 | 3,150 | 13 | 3,076 | 12 |
| | every 7d | 3,025 | 13 | 2,938 | 12 | 2,853 | 11 |
| | every 3 d | 2,195 | 9 | 2,097 | 8 | 1,995 | 8 |
| | every 2d | 1,485 | 6 | 1,414 | 5 | 1,332 | 5 |
| | every 1d | 599 | 3 | 578 | 3 | 546 | 3 |
| $V - S_P$ | every 14d | 3,255 | 14 | 3,180 | 13 | 3,111 | 13 |
| | every 7d | 3,126 | 13 | 3,046 | 12 | 2,971 | 12 |
| | every 3d | 2,683 | 10 | 2,598 | 10 | 2,514 | 9 |
| | every 2d | 2,261 | 8 | 2,181 | 8 | 2,099 | 7 |
| | every 1d | 1,303 | 5 | 1,264 | 5 | 1,212 | 5 |
| $V - \overline{S}$ | N/A | 3,350 | 15 | 3,279 | 14 | 3,214 | 13 |
| **No vaccination** | | | | | | | |
| $\overline{V} - \overline{S}$ | N/A | 3,512 | 16 | 3,442 | 15 | 3,382 | 14 |
| $\overline{V} - S_F$ | every 14d | 3,427 | 15 | 3,357 | 14 | 3,292 | 13 |
| | every 7d | 3,275 | 14 | 3,199 | 13 | 3,123 | 12 |
| | every 3d | 2,561 | 10 | 2,468 | 9 | 2,371 | 9 |
| | every 2d | 1,815 | 7 | 1,739 | 6 | 1651 | 6 |
| | every 1d | 723 | 3 | 701 | 3 | 664 | 3 |

**Table D:** Total number of infections and peak number of hospitalizations for the 80-day semester for all strategies, implemented at various screening frequencies and a vaccination rate of 60 vaccines/day under the base-case scenario, considering various screening ($\eta$) and vaccination ($\alpha$) compliance rates and an initial campus-wide immunity ($L_s = L_f$) of 60%.

| | | $\alpha = 50\%$ | | $\alpha = 75\%$ | | $\alpha = 95\%$ | |
|---|---|---|---|---|---|---|---|
| Strategy | Test Frequency | Total number of infections | Peak number of hospitalizations | Total number of infections | Peak number of hospitalizations | Total number of infections | Peak number of hospitalizations |
| $\eta = 50\%$ | | | | | | | |
| | every 14d | 1,280 | 6 | 1,011 | 4 | 793 | 3 |
| | every 7d | 1,105 | 5 | 848 | 4 | 660 | 2 |
| $V - S_F$ | every 3 d | 732 | 4 | 543 | 3 | 424 | 2 |
| | every 2d | 532 | 3 | 392 | 2 | 310 | 2 |
| | every 1d | 279 | 2 | 203 | 2 | 163 | 1 |
| | every 14d | 1,340 | 6 | 1,077 | 4 | 852 | 3 |
| | every 7d | 1,226 | 6 | 970 | 4 | 765 | 3 |
| $V - S_P$ | every 3d | 979 | 5 | 756 | 3 | 598 | 2 |
| | every 2d | 823 | 4 | 633 | 3 | 495 | 2 |
| | every 1d | 561 | 3 | 429 | 2 | 344 | 2 |
| $\eta = 75\%$ | | | | | | | |
| | every 14d | 1,191 | 6 | 924 | 4 | 720 | 3 |
| | every 7d | 945 | 5 | 711 | 3 | 551 | 2 |
| $V - S_F$ | every 3 d | 532 | 3 | 392 | 2 | 310 | 2 |
| | every 2d | 370 | 3 | 269 | 2 | 216 | 2 |
| | every 1d | 186 | 2 | 135 | 2 | 107 | 1 |
| | every 14d | 1,282 | 6 | 1,024 | 4 | 802 | 3 |
| | every 7d | 1,123 | 5 | 880 | 4 | 689 | 2 |
| $V - S_P$ | every 3d | 823 | 4 | 633 | 3 | 495 | 2 |
| | every 2d | 666 | 3 | 513 | 2 | 400 | 2 |
| | every 1d | 441 | 3 | 337 | 2 | 269 | 1 |
| $\eta = 95\%$ | | | | | | | |
| | every 14d | 1,124 | 5 | 863 | 4 | 671 | 2 |
| | every 7d | 834 | 4 | 623 | 3 | 485 | 2 |
| $V - S_F$ | every 3 d | 434 | 3 | 316 | 2 | 251 | 2 |
| | every 2d | 294 | 2 | 213 | 2 | 171 | 1 |
| | every 1d | 146 | 2 | 104 | 1 | 83 | 1 |
| | every 14d | 1,240 | 6 | 980 | 4 | 772 | 3 |
| | every 7d | 1,051 | 5 | 814 | 3 | 642 | 2 |
| $V - S_P$ | every 3d | 728 | 4 | 559 | 3 | 439 | 2 |
| | every 2d | 577 | 3 | 445 | 2 | 348 | 2 |
| | every 1d | 380 | 2 | 296 | 2 | 233 | 1 |
| **No screening** | | | | | | | |
| $V - \overline{S}$ | N/A | 1,451 | 7 | 1,200 | 5 | 952 | 3 |

**Table E:** Total number of infections and peak number of hospitalizations for the 80-day semester for all strategies, implemented at various screening frequencies and vaccination rates, under three severity scenarios, considering 75% compliance ($\eta = \alpha = 75\%$) and 60% initial campus-wide immunity ($L_s = L_f = 60\%$).

| | | Base-case Scenario | | Best-case Scenario | | Worst-case Scenario | |
|---|---|---|---|---|---|---|---|
| Strategy | Test frequency | Total number of infections | Peak number of hospitalizations | Total number of infections | Peak number of hospitalizations | Total number of infections | Peak number of hospitalizations |
| **120 vaccines/day** | | | | | | | |
| $V - S_F$ | every 14d | 347 | 1 | 149 | 0 | 812 | 2 |
| | every 7d | 281 | 1 | 114 | 0 | 712 | 2 |
| | every 3 d | 171 | 1 | 68 | 0 | 498 | 2 |
| | every 2d | 125 | 1 | 48 | 0 | 387 | 1 |
| | every 1d | 65 | 1 | 25 | 0 | 220 | 1 |
| $V - S_P$ | every 14d | 381 | 1 | 165 | 0 | 860 | 2 |
| | every 7d | 336 | 1 | 137 | 0 | 800 | 2 |
| | every 3d | 250 | 1 | 100 | 0 | 657 | 2 |
| | every 2d | 207 | 1 | 80 | 0 | 561 | 1 |
| | every 1d | 143 | 1 | 53 | 0 | 420 | 1 |
| $V - \overline{S}$ | N/A | 448 | 1 | 196 | 1 | 947 | 2 |
| **60 vaccines/day** | | | | | | | |
| $V - S_F$ | every 14d | 924 | 4 | 438 | 2 | 1,429 | 7 |
| | every 7d | 711 | 3 | 304 | 1 | 1,290 | 7 |
| | every 3 d | 392 | 2 | 152 | 1 | 944 | 5 |
| | every 2d | 269 | 2 | 102 | 1 | 732 | 5 |
| | every 1d | 135 | 2 | 50 | 1 | 406 | 4 |
| $V - S_P$ | every 14d | 1,024 | 4 | 510 | 2 | 1,475 | 8 |
| | every 7d | 880 | 4 | 411 | 2 | 1,394 | 7 |
| | every 3d | 633 | 3 | 262 | 1 | 1,193 | 6 |
| | every 2d | 513 | 2 | 203 | 1 | 1,051 | 5 |
| | every 1d | 337 | 2 | 124 | 1 | 803 | 4 |
| $V - \overline{S}$ | N/A | 1,200 | 5 | 662 | 2 | 1,549 | 8 |
| **30 vaccines/day** | | | | | | | |
| $V - S_F$ | every 14d | 1,323 | 6 | 800 | 3 | 1,616 | 9 |
| | every 7d | 1,708 | 5 | 546 | 2 | 1,521 | 9 |
| | every 3 d | 635 | 3 | 259 | 1 | 1,216 | 8 |
| | every 2d | 443 | 3 | 170 | 1 | 1,003 | 7 |
| | every 1d | 224 | 2 | 83 | 1 | 608 | 5 |
| $V - S_P$ | every 14d | 1,399 | 6 | 908 | 3 | 1,636 | 9 |
| | every 7d | 1,237 | 5 | 710 | 3 | 1,566 | 9 |
| | every 3d | 914 | 4 | 420 | 2 | 1,397 | 8 |
| | every 2d | 735 | 4 | 305 | 2 | 1,258 | 8 |
| | every 1d | 484 | 3 | 173 | 1 | 982 | 7 |
| $V - \overline{S}$ | N/A | 1,561 | 7 | 1,169 | 4 | 1,694 | 10 |
| **No vaccination** | | | | | | | |
| $\overline{V} - \overline{S}$ | N/A | 1,834 | 8 | 1,595 | 6 | 1,828 | 10 |
| $\overline{V} - S_F$ | every 14d | 1,644 | 7 | 1,181 | 4 | 1,777 | 10 |
| | every 7d | 1,408 | 6 | 819 | 3 | 1,715 | 9 |
| | every 3d | 865 | 4 | 364 | 2 | 1,491 | 8 |
| | every 2d | 599 | 3 | 232 | 1 | 1,280 | 7 |
| | every 1d | 294 | 2 | 108 | 1 | 827 | 6 |

**Table F:** Total number of infections and peak number of hospitalizations for the 80-day semester for some strategies, implemented at various screening frequencies and a vaccination rate of 120 vaccines/day, under two severity scenarios, considering 75% compliance ($\eta = \alpha = 75\%$) and 30% initial campus-wide immunity ($L_s = L_f = 30\%$).

| | | Base-case Scenario | | Worst-case scenario | |
|---|---|---|---|---|---|
| Strategy | Test Frequency | Total number of infections | Peak number of hospitalizations | Total number of infections | Peak number of hospitalizations |
| **120 vaccines/day** | | | | | |
| $V - S_F$ | every 14d | 2,419 | 7 | 2,940 | 10 |
| | every 7d | 2,019 | 6 | 2,783 | 10 |
| | every 3 d | 1,121 | 4 | 2,264 | 8 |
| | every 2d | 695 | 3 | 1,773 | 7 |
| | every 1d | 275 | 2 | 895 | 5 |
| $V - S_P$ | every 14d | 2,564 | 8 | 2,987 | 11 |
| | every 7d | 2,348 | 7 | 2,909 | 10 |
| | every 3d | 1,844 | 5 | 2,682 | 8 |
| | every 2d | 1,532 | 4 | 2,479 | 7 |
| | every 1d | 1,024 | 3 | 2,040 | 6 |
| $V - \overline{S}$ | N/A | 2,773 | 9 | 3,059 | 11 |

**Table G:** Total number of infections and peak number of hospitalizations for the 80-day semester for some strategies, implemented at various screening frequencies and a vaccination rate of 120 vaccines/day, under two severity scenarios, considering 50% compliance ($\eta = \alpha = 50\%$) and 30% initial campus-wide immunity ($L_s = L_f = 30\%$).

| | | Base-case Scenario | | Worst-case scenario | |
|---|---|---|---|---|---|
| Strategy | Test Frequency | Total number of infections | Peak number of hospitalizations | Total number of infections | Peak number of hospitalizations |
| **120 vaccines/day** | | | | | |
| $V - S_F$ | every 14d | 2,940 | 13 | 3,158 | 15 |
| | every 7d | 2,755 | 12 | 3,096 | 14 |
| | every 3 d | 2,117 | 9 | 2,880 | 14 |
| | every 2d | 1,559 | 7 | 2,605 | 12 |
| | every 1d | 662 | 4 | 1,730 | 9 |
| $V - S_P$ | every 14d | 2,989 | 13 | 3,170 | 15 |
| | every 7d | 2,886 | 12 | 3,131 | 15 |
| | every 3d | 2,592 | 10 | 3,014 | 13 |
| | every 2d | 2,341 | 9 | 2,895 | 12 |
| | every 1d | 1,775 | 6 | 2,589 | 11 |

**Fig B:** Peak number of hospitalizations versus total number of infections for 60% initial faculty and students vaccination coverage ($L_f = L_s = 60\%$), 75% screening and vaccination compliance rates ($\eta = \alpha = 75\%$) and different severity scenarios, i,e.: (a) base-case, (b) worst-case and (c) best-case (assuming vaccinated individuals are not in the routine screening pool). (d) is a zoomed in version of the best-case scenario. (Order of a, b, c and d goes from left to right and from up to down)

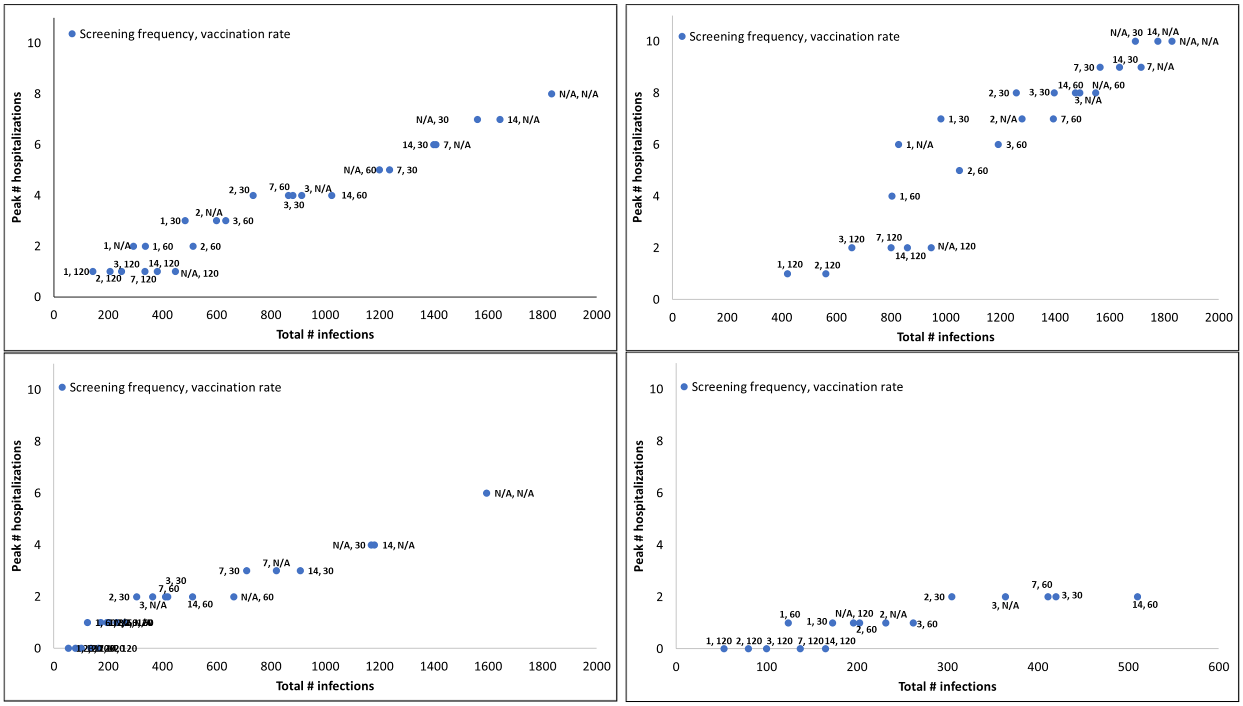

**Fig C:** Peak number of hospitalizations versus total number of infections for the base-case scenario, $\eta = \alpha = 75\%$, 30% initial faculty vaccination coverage ($L_f$) and different initial student vaccination coverage ($L_s$), i,e.: (a) 30%, (b) 60% and (c) 90% assuming vaccinated individuals can get screened. (d) is a zoomed in version of the 90% case. (Order of a, b, c and d goes from left to right and from up to down)

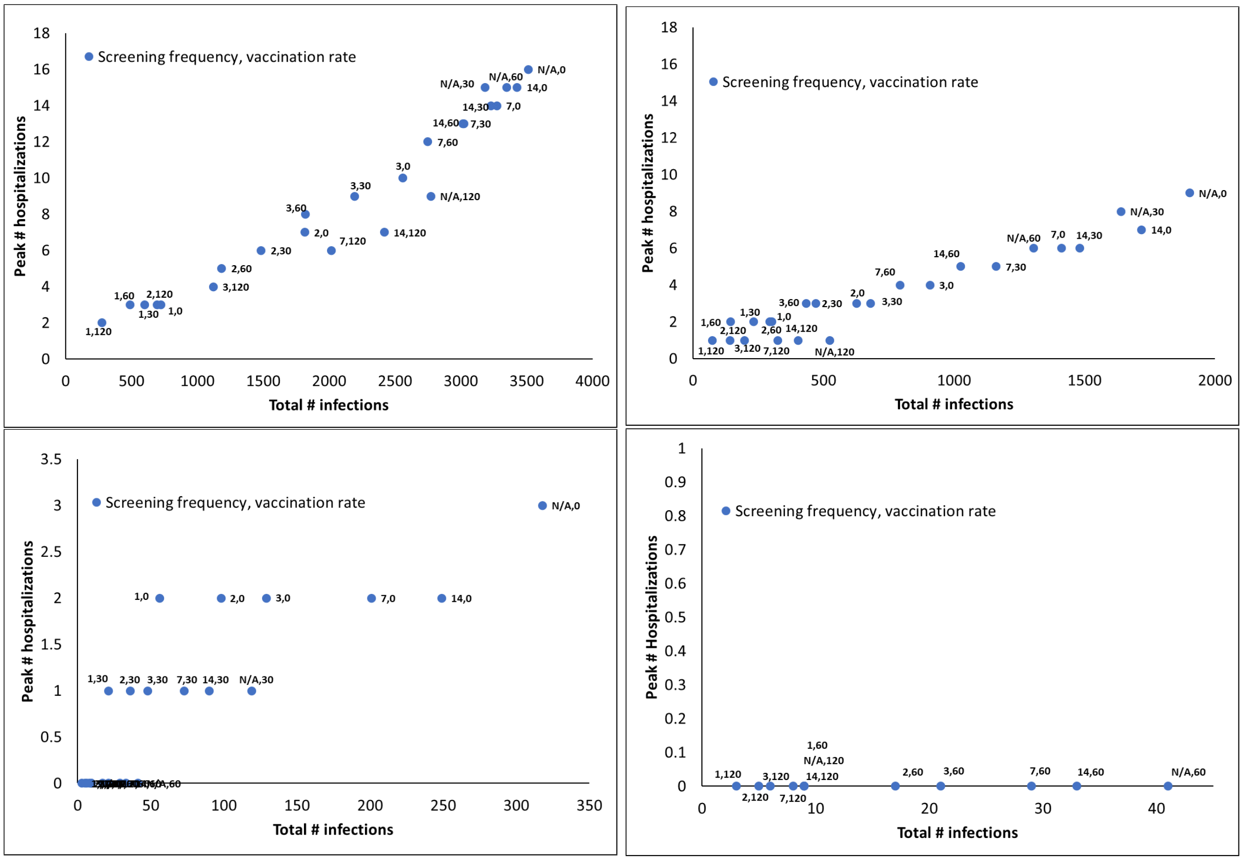

**Fig D:** Peak number of hospitalizations versus total number of infections for the base-case scenario, $\eta = \alpha = 75\%$, 30% initial student vaccination coverage ($L_s$) and different initial faculty vaccination coverage ($L_f$), i,e.: (a) 30%, (b) 60% and (c) 90% assuming vaccinated individuals can get screened. (Order of a, b and c goes from up to down)

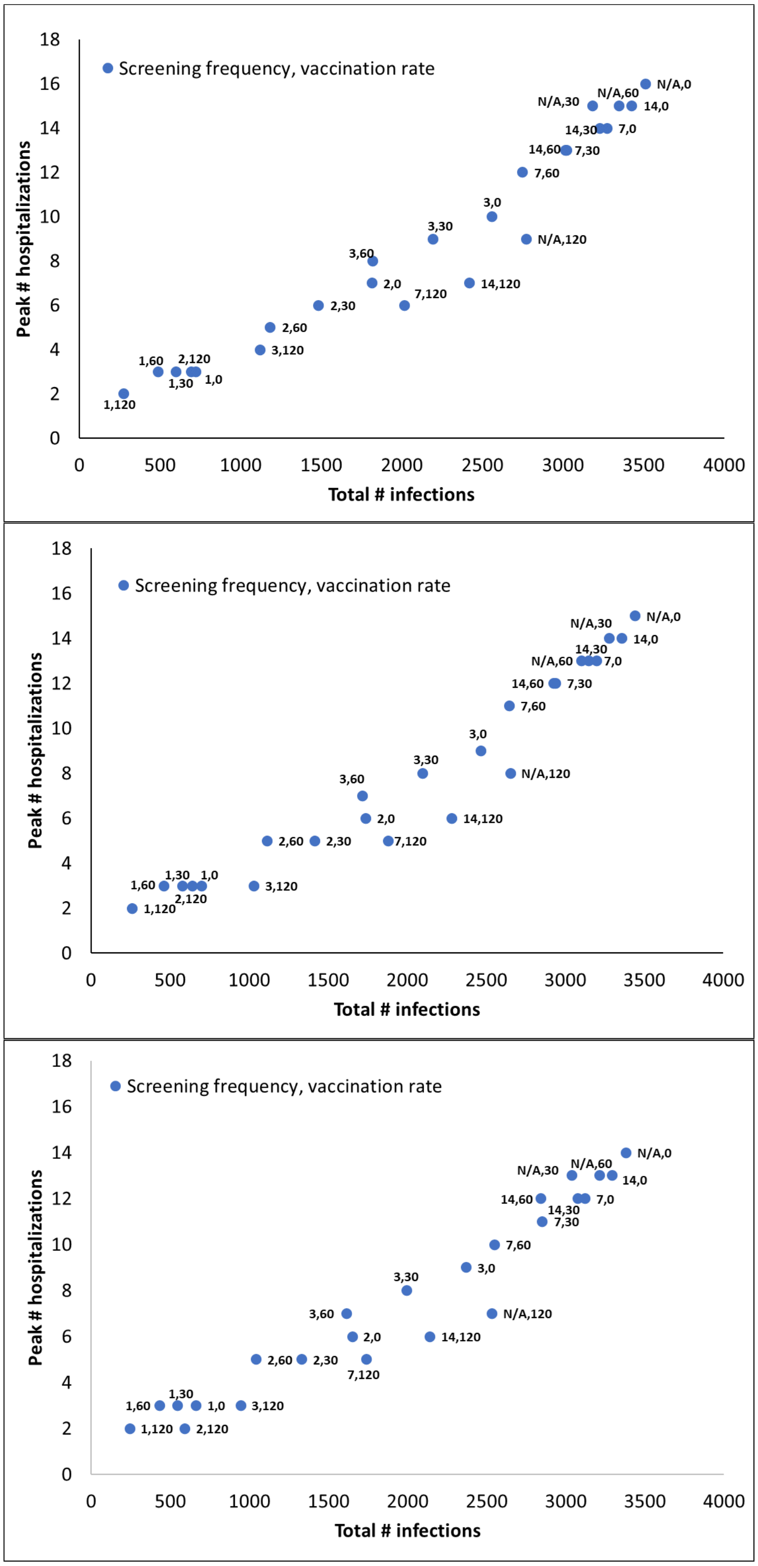

[3] Niche. William & Mary; Accessed on December 2021.

[4] COVID-Net. Laboratory-Confirmed COVID-19-Associated Hospitalizations; Accessed on November 2021.

[5] CDC. Demographic Trends of COVID-19 cases and deaths in the US reported to CDC; Accessed on November 2021.

[6] McChesney J, Bichsel J. The Aging of Tenure-Track Faculty in Higher Education: Implications for Succession and Diversity. College and University Professional Association for Human Resources. 2020;.

[7] Statista. Number of coronavirus disease 2019 (COVID-19) deaths in the U.S. as of November 24, 2021, by age*; Accessed on November 2021.

[8] Statista. Total number of cases of COVID-19 in the United States as of November 24, 2021, by age group; Accessed on November 2021.

[9] Statista. Number of COVID-19 vaccine doses administered in the United States as of November 17, 2021, by vaccine manufacturer; Accessed on November 2021.

[10] CDC. Pfizer-BioNTech COVID-19 Vaccine Overview and Safety (also known as COMIRNATY); Accessed on November 2021.

[11] CDC. Moderna COVID-19 Vaccine Overview and Safety; Accessed on November 2021.

[12] Centers for Disease Control. Different COVID-19 vaccines; Accessed on November 2021.
